# BRSET: A Brazilian Multilabel Ophthalmological Dataset of Retina Fundus Photos

**DOI:** 10.1101/2024.01.23.24301660

**Authors:** Luis Filipe Nakayama, David Restrepo, João Matos, Lucas Zago Ribeiro, Fernando Korn Malerbi, Leo Anthony Celi, Caio Saito Regatieri

## Abstract

**Introduction:** The Brazilian Multilabel Ophthalmological Dataset (BRSET) addresses the scarcity of publicly available ophthalmological datasets in Latin America. BRSET comprises 16,266 color fundus retinal photos from 8,524 Brazilian patients, aiming to enhance data representativeness, serving as a research and teaching tool. It contains sociodemographic information, enabling investigations into differential model performance across demographic groups.

**Methods:** Data from three São Paulo outpatient centers yielded demographic and medical information from electronic records, including nationality, age, sex, clinical history, insulin use, and duration of diabetes diagnosis. A retinal specialist labeled images for anatomical features (optic disc, blood vessels, macula), quality control (focus, illumination, image field, artifacts), and pathologies (e.g., diabetic retinopathy). Diabetic retinopathy was graded using International Clinic Diabetic Retinopathy and Scottish Diabetic Retinopathy Grading. Validation used Dino V2 Base for feature extraction, with 70% training and 30% testing subsets. Support Vector Machines (SVM) and Logistic Regression (LR) were employed with weighted training. Performance metrics included area under the receiver operating curve (AUC) and Macro F1-score.

**Results:** BRSET comprises 65.1% Canon CR2 and 34.9% Nikon NF5050 images. 61.8% of the patients are female, and the average age is 57.6 years. Diabetic retinopathy affected 15.8% of patients, across a spectrum of disease severity. Anatomically, 20.2% showed abnormal optic discs, 4.9% abnormal blood vessels, and 28.8% abnormal macula. Models were trained on BRSET in three prediction tasks: “diabetes diagnosis”; “sex classification”; and “diabetic retinopathy diagnosis”.

**Discussion:** BRSET is the first multilabel ophthalmological dataset in Brazil and Latin America. It provides an opportunity for investigating model biases by evaluating performance across demographic groups. The model performance of three prediction tasks demonstrates the value of the dataset for external validation and for teaching medical computer vision to learners in Latin America using locally relevant data sources.

**Author Summary:** In low-resource settings, access to open medical datasets is crucial for research. Regions such as Latin America often face underrepresentation, resulting in health biases and inequities. To face the scarcity of diverse ophthalmological datasets in these areas, especially in Brazil and Latin America, we introduce the Brazilian Multilabel Ophthalmological Dataset (BRSET) as a means to alleviate biases in medical AI research. Comprising 16,266 color fundus retinal photos from 8,524 Brazilian patients, BRSET integrates sociodemographic information, empowering researchers to investigate biases across demographic groups and diseases. BRSET was extracted from São Paulo outpatient centers, and includes demographics, clinical history, and retinal images labeled for anatomical features, quality control, and pathologies like diabetic retinopathy. Validation was performed in a set of selected prediction tasks, such as diabetes diagnosis, sex classification, and diabetic retinopathy diagnosis. BRSET’s inclusion of sociodemographic data and experiment metrics underscores its potential efficacy across diverse classification objectives and patient groups, providing crucial insights for medical AI in underrepresented regions.

## Introduction

In ophthalmological practice, imaging assists in the diagnosis and follow-up of ocular conditions, including retinal fundus photos, ocular anterior segment photos, corneal topography, visual field tests, and optical coherence tomography [1,2]. Artificial intelligence (AI) algorithms can potentially improve medical care by facilitating access to screening, diagnosis, and monitoring in resource-limited settings and assist with the decision-making process [1–3]. In ophthalmology, AI holds promise for ocular diseases such as diabetic retinopathy, age-related macular degeneration, glaucoma, and retinopathy of prematurity [1,2,4–8]. While AI represents a breakthrough technology, concerns for unfair algorithms resulting from non-representative data and biased models cannot be ignored [9–11].

The open science movement in healthcare has not gained traction in Latin America [14]. In ophthalmology, the majority of the available datasets come from high-income countries, as can be seen in Table 1. In addition, datasets lack demographic and crucial clinical information such as comorbidities [12]. In low and middle-income countries (LMIC), the number of ophthalmologists relative to the population is not adequate [15]. Autonomous systems may increase ophthalmological coverage and reduce preventable blindness; however, datasets that do not adequately represent those who are disproportionately impacted by the disease lead to biased and harmful algorithms [12].

**Table 1:** Comparative table with open-access ophthalmological datasets.

| Dataset | View Position | Labels | Dataset Size | Image format | Annotation level | Retina specialist | Anatomical label | Diabetic retinopathy classification | Nationality |
| --- | --- | --- | --- | --- | --- | --- | --- | --- | --- |
| Eye Picture Archive Communication System | Macular | Diabetic Retinopathy | 88702 images | JPG | Global | NA | No | ICDR | USA |
| BRSET | Macular | Multiple | 16266 images of 8524 patients | JPG | Global | Yes | Yes | ICDR and SDRG | Brazil |
| Jichi DR | Macular | Diabetic Retinopathy | 9939 images | JPG | Global | NA | No | Davis | Japan |
| Asian Pacific Tele Ophthalmology society dataset | Macular | Diabetic Retinopathy | 5593 images | PNG | Global | NA | No | ICDR | India |
| Ocular Disease Recognition (ODIR) | Macular | Normal, diabetes, glaucoma, cataract, amd, hypertension, pathological myopia, other | 5000 images of 5000 patients | JPG | Global | NA | No | No | China |
| Retinal Fundus Multi-Disease Image Dataset (RFMiD): A Dataset for Multi-Disease Detection Research | Macula and Optic disc | Multiple | 3200 images | PNG | Global | NA | No | No | India |
| Messidor 2 | Macular | Diabetic Retinopathy | 1748 images | JPG, PNG | Global | No | No | No | France |
| DR1/DR2 | Macular | Diabetic Retinopathy | 1597 images | TIFF | Segmentation | NA | No | No | Brazil |
| Rotterdam Ophthalmic data repository | Macular and mosaic | Diabetic Retinopathy | 1120 images | PNG | Global | NA | No | No | Netherlands |
| Indian Diabetic Retinopathy Image Dataset | Macular | Diabetic Retinopathy | 516 images | JPG | Global and Segmentation | Yes | No | ICDR | India |
| DIARETDB0 | Macular | Diabetic Retinopathy | 130 images | PNG | Segmentation | NA | No | No | Finland |
| DIARETDB1 | Macular | Diabetic Retinopathy | 89 images | PNG | Segmentation | Yes | No | No | Finland |
| E-ophta | Macular | Diabetic Retinopathy | 463 images | JPG | Segmentation | NA | No | No | France |
| Hamilton Eye Institute Macular Edema | Macular | Diabetic Retinopathy | 169 images | JPG | Segmentation | Yes | No | No | USA |

BRSET is the first publicly available Latin American ophthalmological dataset across a range of disease severity and sociodemographic categories.

## Materials and methods

This study was approved by the Sao Paulo Federal University (UNIFESP) IRB (CAAE 33842220.7.0000.5505) and included retinal fundus photos and clinical data. In this dataset, identifiable patient information was removed from all images.

### Data sources

We included data from three outpatient Brazilian ophthalmological centers in São Paulo evaluated from 2010 to 2020 and from the Sao Paulo Federal University ophthalmology sector.

### Data Collection

Images present in this dataset were collected using different retinal fundus cameras, including Nikon NF505 (Nikon, Tokyo, Japan), Canon CR-2 (Canon Inc, Melville, NY, USA) retinal camera, and Phelcom Eyer (Phelcom Technologies, MA, USA). Retinal photos were taken by previously trained non-medical professionals in pharmacological mydriasis.

### Dataset preparation

The file identification was removed from all fundus photos, as well as sensitive data (e.g., patient name, exam date). Every image was reviewed to ensure the absence of protected health information in images. The images were exported directly from retinal cameras in JPEG format, and no preprocessing techniques were performed. The image viewpoint can be macula-centered or optic disc-centered. The dataset does not include fluorescein angiogram photos, non-retinal images, or duplicated images.

### Metadata

Each retinal image is labeled with the retinal camera device, image center position, patient nationality, age in years, sex, comorbidities, insulin use, and duration of diabetes diagnosis. The demographics and medical features were collected from the electronic medical records.

### Labeling

A retinal specialist ophthalmologist labeled all the images based on criteria that were established by the research group [16]. The following characteristics were labeled:

#### Anatomic classification

The retinal optic disc (vertical cup-disc ratio of ≥0.65 [17]), retinal vessels (tortuosity and width), and macula (abnormal findings) were categorized as either normal or abnormal.

#### Quality control parameters

Parameters including image focus, illumination, image field, and artifacts, were assessed and classified as satisfactory or unsatisfactory. The criteria are defined in Table 2.

**Table 2:** Quality assessment parameters.

|  |  |
| --- | --- |
| **Illumination** | This parameter is graded as adequate when both of the following requirements are met: 1) Absence of dark, bright, or washed-out areas that interfere with detailed grading; 2) In the case of peripheral shadows (e.g., due to pupillary constriction) the readable part should reach more than 80% of the whole image. |
| **Image Field** | This parameter is graded as adequate when all the following requirements are met: 1) The optic disc is at least 1 disc diameter (DD) from the nasal edge; 2) The macular center is at least 2 DD from the temporal edge; 3) The superior and inferior temporal arcades are visible in a length of at least 2 DD |
| **Artifacts** | The following artifacts are considered: haze, dust, and dirt. This parameter is graded as adequate when the image is sufficiently artifact-free to allow adequate grading. |
| **Focus** | This parameter is graded as adequate when the focus is sufficient to identify third-generation branches within one optic disc diameter around the macula. |

#### Pathological classifications

The images were classified according to the pathological classification list: diabetic retinopathy, diabetic macular edema, scar (toxoplasmosis), nevus, age-related macular degeneration (AMD), vascular occlusion, hypertensive retinopathy, drusens, nondiabetic retinal hemorrhage, retinal detachment, myopic fundus, increased cup disc ratio, other.

#### Diabetic retinopathy classification

Diabetic retinopathy was classified using the International Clinic Diabetic Retinopathy (ICDR) grading and Scottish Diabetic Retinopathy Grading (SDRG), as can be seen in Table 3 [18,19].

**Table 3:** Diabetic Retinopathy Classifications.

| Classification | 0 - Normal | 1 - Mild non-proliferative DR | 2- Moderate non-proliferative DR | 3 - Severe non-proliferative DR | 4 - Proliferative DR | Macular edema |
| --- | --- | --- | --- | --- | --- | --- |
| **International Classification of** | No abnormalities | Microaneurysms only | More than just microaneurysms but less than severe non-proliferative | Any of the following:<br>> 20 intra-retinal hemorrhages in each of 4 quadrants, definite venous | One or more of the following: neovascularization and/or vitreous or | Exudates or apparent thickening within one disc diameter from the fovea |
| **Diabetic Retinopathy** | | | diabetic retinopathy | beading in $\geq 2$ quadrants, prominent intraretinal microvascular abnormalities in $\geq 1$ quadrant, or no signs of proliferative retinopathy | preretinal hemorrhages and/or panfotocoagulation scars | |
| **Scottish Diabetic Retinopathy Grading** | No abnormalities | At least one microaneurysm, flame exudate, blot hemorrhage with or without hard exudate | More than 4 blot hemorrhages in one hemifield | More than 4 blot hemorrhages in both hemifields, IRMA, venous beading | Disc neovessels, retinal neovessels, vitreous hemorrhage, retinal detachment | Hard exudates within 1-2 DD of the fovea |

### Data Records

This dataset may be used to build computer vision models that predict demographic characteristics and multi-label disease classification. BRSET consists of 16,266 images from 8,524 Brazilian patients, and a metadata file called *labels*.*csv*. Columns are detailed in Table 4.

**Table 4.** Data Dictionary – description of present columns.

| Group | Column Name | Description |
| --- | --- | --- |
| Descriptive and Demographic fields | *image_id* | Image identifier. |
|  | *patient_id* | Patient identifier |
|  | *camera* | Retinal camera (Canon CR or NIKON NF5050). |
|  | *patient_age* | Age of patient in years. |
|  | *comorbidities* | Free text of self-referred clinical antecedents. |
|  | *diabetes_time* | Self-referred time of diabetes diagnosis in years. |
|  | *insulin_use* | Self-referred use of insulin (yes or no). |
|  | *patient_sex* | Enumerated values: 1 for male and 2 for female. |
|  | *exam_eye* | Enumerated values: 1 for the right eye and 2 for the left eye. |
|  | *nationality* | Patient's nationality. |
| Anatomical parameters | *optic_disc* | Enumerated values: 1 for normal and 2 for abnormal. |
|  | *vessels* | Enumerated values: 1 for normal and 2 for abnormal. |
|  | *macula* | Enumerated values: 1 for normal and 2 for abnormal. |
| Diabetes | *diabetes* | Diabetes diagnosis |
| Diabetic retinopathy classification | *DR_ICDR* | International Clinic Diabetic Retinopathy classification with enumerated values from 0 to 4:<br>- 0 No retinopathy.<br>- 1 Mild non-proliferative diabetic retinopathy.<br>- 2 Moderate non-proliferative diabetic retinopathy.<br>- 3 Severe non-proliferative diabetic retinopathy.<br>- 4 Proliferative diabetic retinopathy and post-laser status. |
|  | *DR_SDRG* | Scottish Diabetic Retinopathy Grading Scheme classification with enumerated values from 0 to 4:<br>- 0 No retinopathy.<br>- 1 Mild Background.<br>- 2 Moderate Background.<br>- 3 Severe non-proliferative or pre-proliferative diabetic retinopathy.<br>- 4 Proliferative diabetic retinopathy and post-laser status. |
| Quality parameters | *focus* | Enumerated values: 1 for normal and 2 for abnormal. |
|  | *illumination* | Enumerated values: 1 for normal and 2 for abnormal. |
|  | *image_field* | Enumerated values: 1 for normal and 2 for abnormal. |
|  | *artifacts* | Enumerated values: 1 for normal and 2 for abnormal. |
| Classification parameters | *diabetic_retinopathy* | 1 present and 0 absent. |
|  | *macular_edema* | 1 present and 0 absent. |
|  | *scar* | 1 present and 0 absent. |
|  | *nevus* | 1 present and 0 absent. |
|  | *amd* | 1 present and 0 absent. |
|  | *vascular_occlusion* | 1 present and 0 absent. |
|  | *hypertensive_retinopathy* | 1 present and 0 absent. |
|  | *drusens* | 1 present and 0 absent. |
|  | *hemorrhage* | 1 present and 0 absent. |
|  | *retinal_detachment* | 1 present and 0 absent. |
|  | *myopic_fundus* | 1 present and 0 absent. |
|  | *increased_cup_disc* | 1 present and 0 absent. |
|  | *other* | 1 present and 0 absent. |

### Data storage

The dataset images and labels are stored on the PhysioNet repository entitled “A Brazilian Multilabel Ophthalmological Dataset (BRSET)”. Link: https://physionet.org/content/brazilian-ophthalmological/1.0.0/.

### Descriptive Analytics and Technical Validation

#### Descriptive Analysis

The BRSET database contains 10,592 (65.1%) images taken from a Canon CR2 retinal camera, and the resting 5,674 (34.9%) are from the Nikon NF5050 retinal camera. The sex distributions in the dataset are 6,214 (38.2%) male patients, and 10,052 (61.8%) female patients. The average age is 57.6 years (with a standard deviation of 18.3 years), as can be seen in Figure 1.

**Figure 1.**
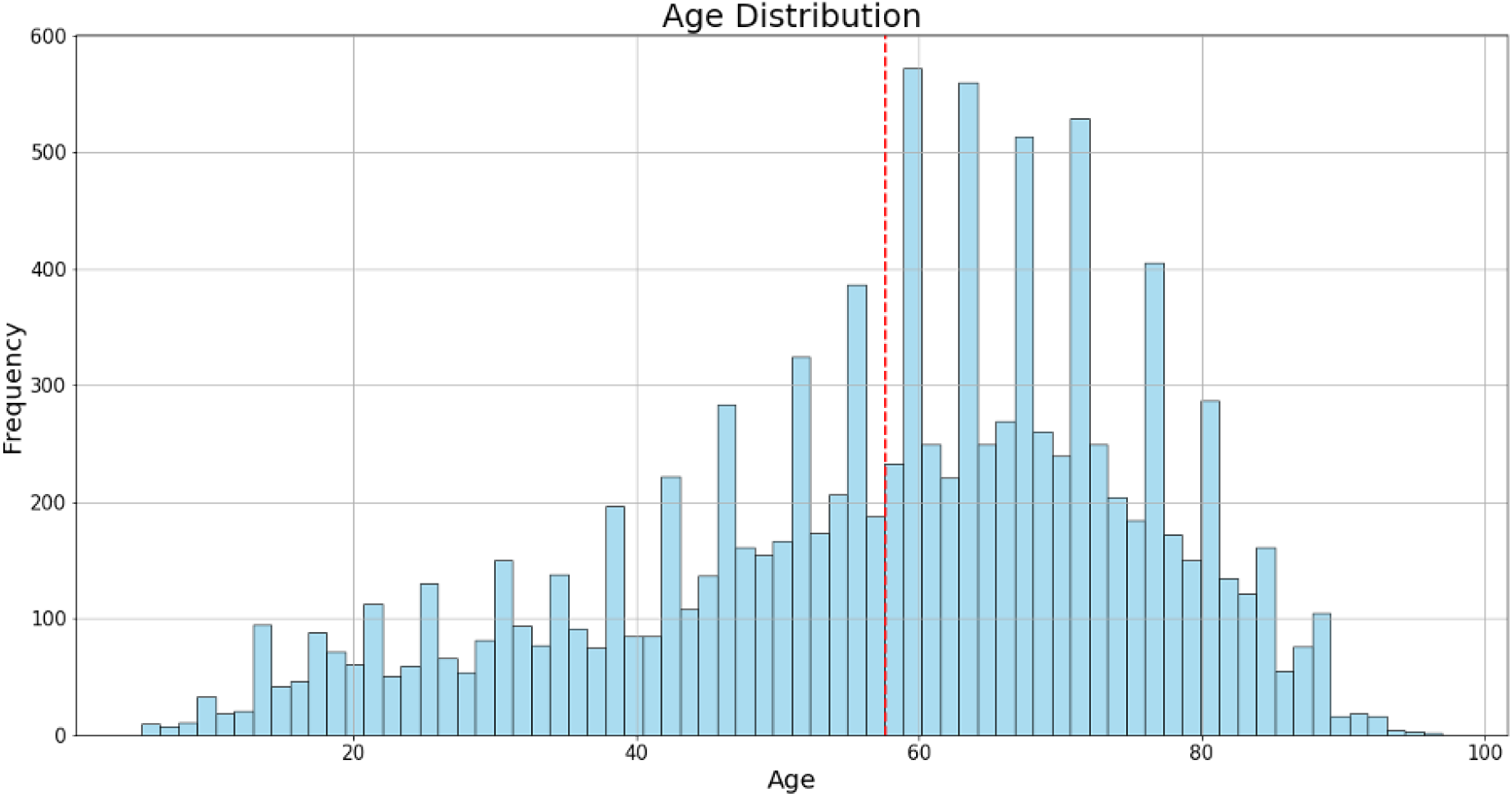
Histogram of age distribution in BRSET. The x-axis represents age groups, while the y-axis indicates the frequency of individuals in each age category.

The database includes 2,579 (15.8%) patients with a diagnosis of diabetes mellitus. Among those patients 1,922 (74.5%) do not have retinopathy, 107 (4.1%) have mild non-proliferative retinopathy, 181 (7%) have moderate non-proliferative retinopathy, 127 (4.9%) have severe non-proliferative retinopathy, and 242 (9.4%) have proliferative retinopathy. A sample of the images for patients with and without diabetic retinopathy can be seen in Figure 2.

**Figure 2.**
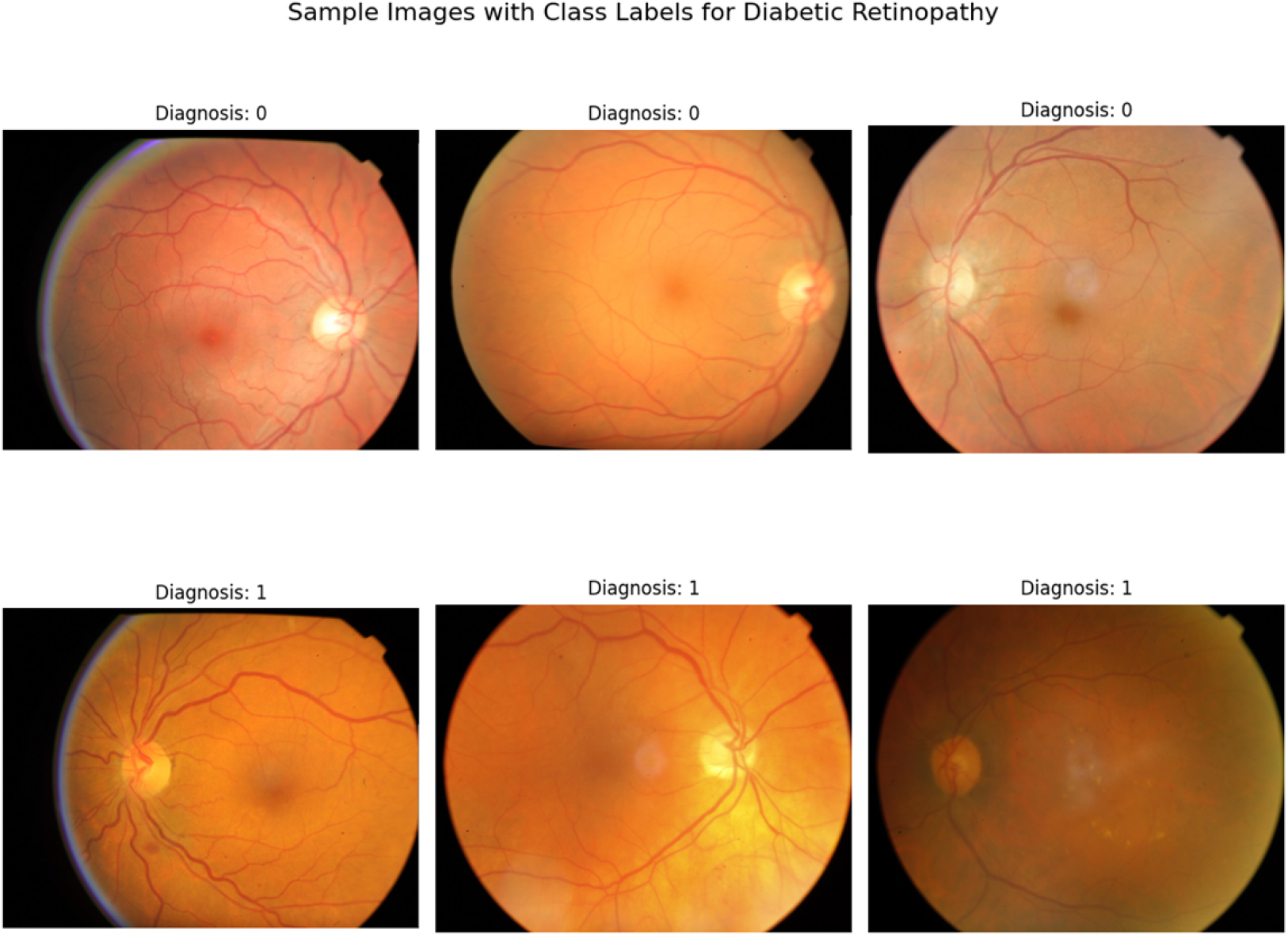
Sample Retina Images with and without Diabetic Retinopathy (DR) from the BRSET Dataset. This figure presents a visual sample of three six images selected from the dataset, with labels indicating the presence (1) or absence (0) of diabetic retinopathy (DR).

Among the anatomical criteria, 3,281 (20.2%) of the images have an abnormal optic disc, 807 (4.9%) have abnormal blood vessels, and 4,685 (28.8%) have an abnormal macula. The distribution of normal exams and pathological findings is described in Table 5.

**Table 5:** Distribution of retina images according to diagnoses and anatomical criteria.

| Labels | Images (n) | Percentage (%) |
| --- | --- | --- |
| Normal | 8,460 | 52 |
| Diabetic retinopathy | 1,046 | 6.4 |
| Diabetic macular edema | 402 | 2.5 |
| Scar | 290 | 1.8 |
| Nevus | 134 | 0.8 |
| Age-related macular degeneration | 366 | 2.2 |
| Vascular occlusion | 103 | 0.6 |
| Hypertensive retinopathy | 283 | 1.7 |
| Drusens | 2,807 | 17.2 |
| Hemorrhage | 96 | 0.6 |
| Retinal detachment | 7 | 0.04 |
| Myopic fundus | 268 | 1.6 |
| Increased cup-disc ratio | 3,202 | 19.7 |
| Other | 758 | 4.7 |

#### Data Quality Assessment

##### Image Quality

In terms of quality criteria, 542 (3.3%) images were classified as having inadequate focus, 84 (0.5%) as having inadequate lighting, 1401 (8.6%) as having an inadequate field, and 57 (0.3%) as having artifacts.

##### Quality Metadata

On the other hand, regarding the quality characteristics of the metadata duplicate data and missing data were assessed. The dataset does not have duplicate data, and only 3 columns contain missing data. As can be seen in Figure 3, the columns patient_age, diabetes_time_y, and insulin contain missing values.

**Figure 3.**
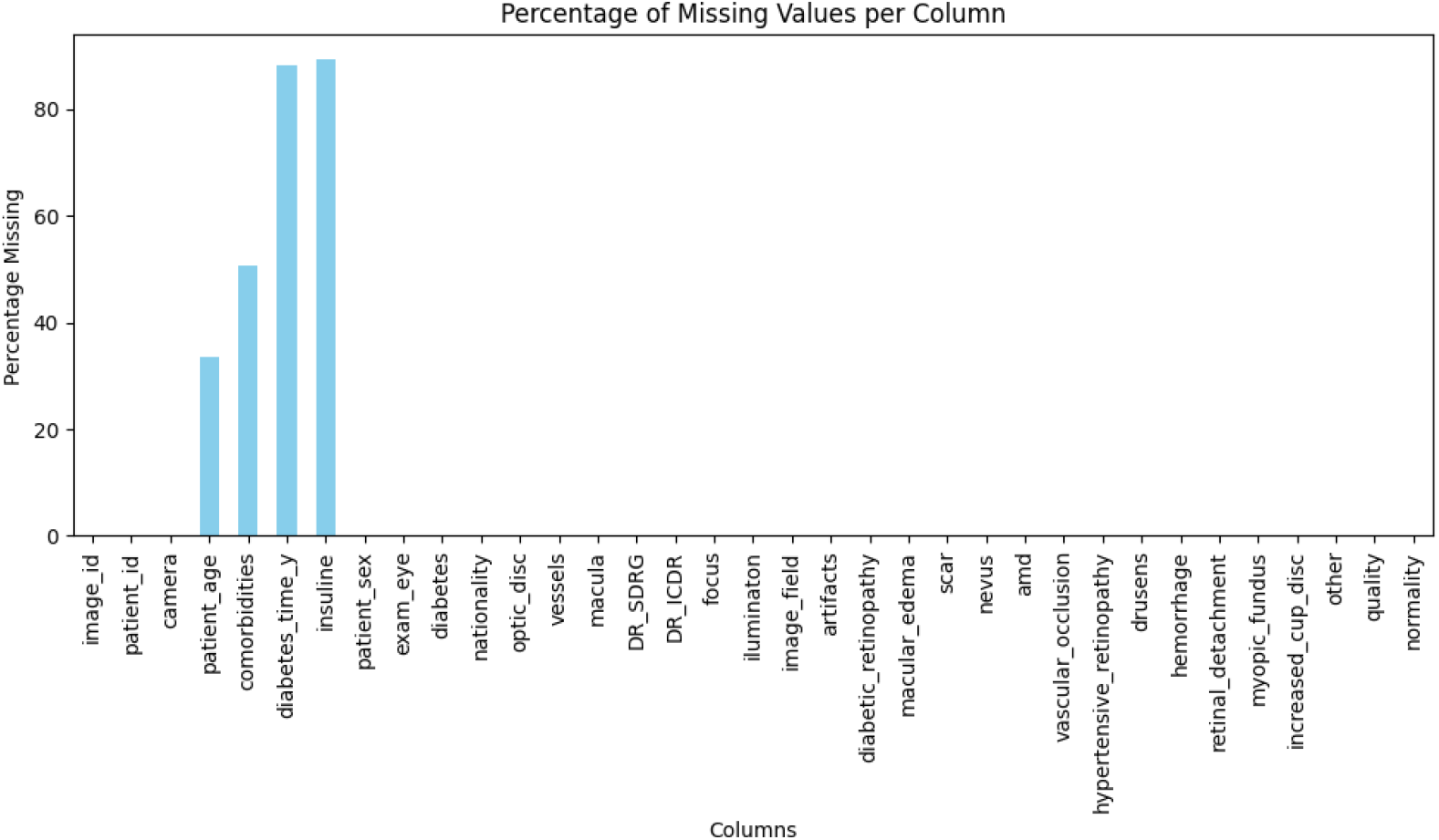
Missing Values Percentage per Column in the BRSET Dataset.

##### Experiments

To explore the dataset, we provide use cases and benchmarks for future research. The codes required to run these experiments are available in a publicly available GitHub repository (see Code availability section).

Dino V2 Base [18], a foundational computer vision model, was used in all the experiments to extract embeddings from the BRSET images. Embeddings are representations of the original data in a lower dimensionality space, which allows us to develop a parsimonious solution, without the need for large computational resources. Although the use of these embeddings provides an effective and computationally efficient strategy, these results must be interpreted as a baseline, and other models or more complex techniques are likely to improve the tasks’ performance.

After being extracted, the image embeddings were saved in a CSV file with 769 columns, where the first column represented the identifier of the image, and the remaining 768 columns represented the embeddings of each image. These resulting embeddings were used as features to train classification Machine Learning (ML) models. The three tasks were binary classification tasks: “diabetes diagnosis”, “sex classification”, and “diabetic retinopathy diagnosis”.

The dataset was divided into training and testing using 70% (11,386 embeddings) for training and 30% (4,880 embeddings) for evaluation. The chosen models for the classification tasks were Support Vector Machines (SVM) with linear kernel and Logistic Regression (LR). The models were trained using class weights to tackle class imbalance during training. The weights were calculated using the equation (1), where w are the weights of class C, N is the total samples in the training dataset, K is the number of classes, and Nc is the samples of class in the training dataset. Finally, the AUC, and the Macro F1-score, given the imbalance, were measured as performance metrics (Table 6).

**Table 6:** Performance metrics of the BRSET for the three selected binary classification tasks. The tasks are “diabetes diagnosis”, “sex classification”, and “diabetic retinopathy diagnosis”.

| Task | Model | AUC | F1-score |
| --- | --- | --- | --- |
| Diabetes diagnosis | SVM | 0.77 | 0.62 |
|  | LR | 0.75 | 0.67 |
| Sex classification | SVM | 0.66 | 0.65 |
|  | LR | 0.68 | 0.68 |
| Diabetic retinopathy diagnosis | SVM | 0.86 | 0.75 |
|  | LR | 0.87 | 0.75 |

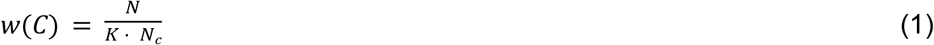

As depicted in Table 6, the dataset can be used, not only for clinical classification tasks but also to identify and quantify differential model performance across sex.

## Usage notes

BRSET is available on PhysioNet as a database that requires user credentialing prior to access. Users must be registered on PhysioNet, have proper human subject research training, and sign a data use agreement that forbids re-identification of patients and sharing it those who are not credentialed.

## Limitations

Our dataset only includes one nationality and represents general ophthalmological outpatients. As a result, the disease distribution is imbalanced, with high percentage of normal and mild cases.

## Strengths

The BRSET is the first multilabel ophthalmological dataset from Brazil and Latin America. BRSET aims to improve data representativeness and create the framework for ophthalmological dataset development. To the best of our knowledge, BRSET is the only publicly available retinal image dataset that also contains sociodemographic data such as sex and age in Latin America, which allows us to investigate algorithmic bias across different demographic groups.

## Code availability

All the codes used in this paper for the dataset setup, data analysis, and experiments are found in a GitHub repository at https://github.com/luisnakayama/BRSET. Best practice guidelines should be followed when analyzing the data, and we incentivize sharing codes and models to promote reproducibility.

## Release note

We plan to include self-declared race and demographic data and increase the ophthalmological exam modalities for future releases.

## Data Availability

All the codes used in this paper for the dataset setup, data analysis, and experiments are found in a GitHub repository at https://github.com/luisnakayama/BRSET. The BRSET is available in https://physionet.org/content/brazilian-ophthalmological/1.0.0/.

## Acknowledgments

LFN was funded by Instituto da Visão - IPEPO and Lemann Foundation.

LAC is funded by the National Institute of Health through R01 EB017205, DS-I Africa U54 TW012043-01 and Bridge2AI OT2OD032701, and the National Science Foundation through ITEST #2148451.

